# Cancer in HIV-positive and HIV-negative adolescents and young adults in South Africa: a cross-sectional study

**DOI:** 10.1101/2020.08.18.20176289

**Authors:** Tafadzwa Dhokotera, Julia Bohlius, Matthias Egger, Adrian Spoerri, Jabulani Ncayiyana, Gita Naidu, Victor Olago, Marcel Zwahlen, Elvira Singh, Mazvita Muchengeti

## Abstract

**Objective:** To determine the spectrum of cancers in AYAs living with HIV in South Africa compared to their HIV negative peers.

**Design:** Cross sectional study with cancer data provided by the National Cancer Registry and HIV data from the National Health Laboratory Service.

**Setting and participants:** The NHLS is the largest provider of pathology services in the South African public sector with an estimated coverage of 80%. The NCR is a division of the NHLS. We included AYAs (aged 10-24 years) diagnosed with cancer by public health sector laboratories between 2004 and 2014 (n=8 479). We included 3 672 in the complete case analysis.

**Primary and secondary outcomes:** We used linked NCR and NHLS data to determine the spectrum of cancers by HIV status in AYAs. We also used multivariable logistic regression to describe the association of cancer in AYAs with HIV, adjusting for age, sex (as appropriate), ethnicity, and calendar period. Due to the large proportion of unknown HIV status we also imputed (post-hoc) the missing HIV status.

**Results:** From 2004-2014, 8 479 AYAs were diagnosed with cancer, HIV status was known for only 45% (n=3812); of those whose status was known, about half were HIV positive (n=1853). AYAs living with HIV were more likely to have Kaposi’s sarcoma (adjusted odds ratio (aOR) 218, 95% CI 89.9-530), cervical cancer (aOR 2.18, 95% CI 1.23-3.89), non-Hodgkin’s lymphoma (aOR 2.12, 95% CI 1.69-2.66), and anogenital cancers other than cervix (aOR 2.73, 95% CI 1.27-5.86). About 44% (n=1 062) of AYAs with HIV related cancers had not been tested for HIV, though they were very likely to have the disease.

**Conclusions:** Cancer burden in AYAs living with HIV in South Africa could be reduced by screening young women for cervical cancer and vaccinating them against human papilloma virus (HPV) infection.

**Strength and limitations:**

- This is the first nationwide study in South Africa to compare the distribution of cancers in adolescents and young adults (AYAs) by HIV status.
- The record linkage and the additional results determined from the text mining process ensured that we extracted the maximum available HIV results.
- We assumed a CD4 count test indicates being HIV positive but CD4 testing maybe performed for other reasons
- Since this was a population of only AYAs diagnosed with cancer, the odds ratios could be overestimated or underestimated depending on the frequency of the cancer

## INTRODUCTION

In Eastern and Southern Africa, an estimated 1.1 million adolescents aged 10-19 years are living with HIV.[1] Young people aged 15-25 years currently make up 30% of new infections.[2] Children infected with HIV perinatally are now more likely to live and to become adolescents and young adults (AYA).[3,4] The outcomes of AYAs living with HIV (AYALHIV) have been poor, mainly because it is challenging to retain them in care. They also tend to have poor virologic suppression, and their CD4 counts often drop to levels that endanger their health.[3,5–7] Co-infection with other oncogenic viruses is also common in this age group.[8,9] For those living with HIV, immunodeficiency and co-infections with other oncogenic viruses are risk factors for developing malignancies. Still, data that compare cancer risk in AYALHIV to that of their HIV negative peers in the antiretroviral therapy era is scarce in resource-limited settings.

Several studies have shown that the risk of HIV-related cancers—non-Hodgkin’s lymphoma (NHL), Kaposi’s sarcoma (KS) and cervical cancer (CC) is higher in AYALHIV than in HIV-negative AYA.[10–15] In the USA, the incidence of leiomyosarcoma was also higher in AYALHIV than in their peers from the general population.[10] However, most of the existing data is from settings with a low HIV burden, but we still know little about cancer burden and risk in AYALHIV in high HIV burden African countries, like South Africa.

We aimed to evaluate the spectrum and cancers associated with HIV in AYAs at a national level. The South African HIV Cancer Match (SAM) study was created to identify the risk factors and spectrum of malignancies in people living with HIV based on routine reports.[16] In this cross-sectional sub analysis, we included AYAs with a pathology-confirmed cancer diagnosis. We examined the proportion of cancer diagnoses with or without HIV infection and the risk factors for cancer in AYAs living with HIV.

## METHODS

### Study design and setting

This was a cross-sectional study with cancer data provided by the National Cancer Registry (NCR) and HIV data from the National Health Laboratory Service (NHLS). The NHLS is the largest provider of diagnostic pathology services in the South African public sector (estimated coverage is over 80% of the SA population).[17] The NHLS include the National Institute of Communicable Diseases, the National Institute of Occupational Health, and the pathology-based NCR. The Corporate Data Warehouse (CDW) is the centralised data centre of the NHLS where all the data on tests performed in its laboratories are stored.

### Inclusion criteria

We included all AYAs with a primary incident cancer recorded from 2004 to 2014 in NCR records. Adolescence was defined as 10 to 19 years and young adulthood as 20 to 24 years at the time of cancer diagnosis, based on World Health Organisation (WHO) and South African Department of Health definitions[6,18]. We excluded cancer precursors and only retained laboratory-confirmed cancer records that contained the International Classification of Disease in Oncology version 3 (ICD-O-3) topography and morphology descriptions.[19] If a person had two different cancers at different sites, they were considered as two individual records (multiple primaries).

### Outcome and exposure variables

The main exposure was HIV infection and the main outcome cancer diagnosis by morphological subtype. HIV status was determined from HIV diagnostic tests (ELISA, qualitative PCR and rapid HIV tests) and HIV monitoring tests (CD4 counts and HIV RNA viral loads). We assumed an individual was HIV positive if any diagnostic test was positive or if monitoring tests (CD4 cell count, viral load) were recorded. We used text mining methods to extract additional HIV results from the clinical history section of cancer pathology reports.

We used data from the CDW patient linking process which utilises probabilistic record linkage (PRL) methods to create a unique patient identifier for records belonging to the same person. As described in detail elsewhere[20], the CDW uses names, surnames and date of births as linkage variables, which are fed into the PRL linkage algorithm. First names and surnames have a weight of 40% each, and date of birth a weight of 20%. Records with a recorded national identity number are exact matches. To be considered a match, the cumulative score has to reach 90% or above. The data from the CDW has been evaluated for completeness and accuracy and validated as a good source of data for research on HIV in South Africa.[21]

We used NCR records to determine demographic characteristics. Where missing, the NCR imputes ethnicity based on surnames using known surname-ethnicity pairings.[22] Ethnicity was grouped into Black and Other for comparison purposes because few subjects belong to other population subgroups. We divided calendar years into three periods: the early years of combination antiretroviral therapy (ART) (2004–2008); later years (2009–2011); and, the most recent period (2012–2014). We selected cut-offs for the calendar periods to correspond with changes in ART guidelines in South Africa during the study period.[23] We grouped cancers of vulva, penis, vagina and anus as anogenital cancers other than cervical cancer. We looked at NHLs as a group, and at each of its subtypes: Burkitt lymphoma; diffuse large B cell lymphoma (DLBCL); diffuse immunoblastic large B cell lymphoma (DILBCL); follicular NHL; and, mature T cell NHL.

### Data analysis

For descriptive purposes, we present sex, ethnicity (Asian, Black, Coloured and White) and age strata by HIV status (positive/negative/unknown). We show the frequency and spectrum of cancers in AYAs stratified by HIV status (positive/negative/unknown) and by sex. We used a logistic regression model to determine the association between HIV and cancer in adolescents and young adults. For each cancer, we used records without the cancer under study as the comparison group, including cancers with an infectious aetiology. We adjusted the models using age (adolescence versus (vs) young adults), sex (male vs female, except for sex-specific cancers), ethnicity (Black vs other) and ART era. We restricted our main analysis to cancers in AYAs with known HIV status, so all AYA were either HIV-positive or HIV-negative. We checked for interactions between HIV and the other factors of interest (age, sex, ethnicity and calendar period) and adjusted models for interaction analysis for age, sex, ethnicity and calendar period. To test for significance of the interaction, we used likelihood ratio tests to compare logistic regression models with and without the interaction terms at 5% significance level. Stata^®^ 15.1 was used for all analyses (StataCorp Inc, College Station, TX, USA).

### Sensitivity analysis

As a sensitivity analysis, we used multiple imputation methods to impute missing HIV results for 4431 cancer patients with unknown HIV status. We included HIV status (the primary exposure), cancer diagnosis, cancer diagnosis year, sex, age and ethnicity in our imputation model. Since ethnicity was already imputed by the NCR using surname-ethnicity pairings, we excluded records that still had missing ethnicity data (4%; n=368). We also excluded records with missing sex as they were few (0.09%; n=8). We use multivariate imputation with chained equations to generate 15 imputed datasets that we combined to give a pooled estimate (odds ratio). We fit multivariable logistic regression models adjusting for age, sex, ethnicity and calendar period. We compared the results from the imputed dataset with the main complete case analysis. Table S1 in the supplement shows the distribution of known and unknown HIV status by the variables in the imputation model.

## RESULTS

Over the 11 years, 8 479 AYAs were diagnosed with cancer. Over half (n=4 466) of all recorded cancer cases were YA (20–24 years), median age 20 years (interquartile range (IQR): 15–23). Girls and women made up 54% (n=4 605) of the AYA population; most AYAs with cancer were Black 75% (n=6 376). About 45% (n=3 819) of AYAs with cancer were assigned an HIV status; half of those with known status were HIV positive (n=1 855). When we compared AYA cancer patients with and without HIV, the median age of AYA cancer patients with HIV was 22 years (IQR: 19–23) while it was 18 years in those without HIV (IQR: 13–21). Those with HIV were more often female (67% vs 45%; p-value < 0.001) and more often Black population (86% vs 64%) (Table 1). The proportion of people with unknown HIV status declined across the calendar period (Figure 1).

**Table 1:** Demographic characteristics of Adolescents and Young Adults with a cancer diagnosis stratified by HIV status in the South African Public Health sector, 2004-2014.

|  | HIV negative | HIV positive | HIV unknown |
| --- | --- | --- | --- |
|  | n (%) | n (%) | n (%) |
| **Age** |  |  |  |
| Median age (Interquartile range) [years] | 18 (13-21) | 22 (19-23) | 20 (16-22) |
| 10-14 | 635 (32.3%) | 200 (10.8%) | 922 (19.8%) |
| 15-19 | 585 (29.8%) | 338 (18.2%) | 1331 (28.6%) |
| 20-24 | 744 (37.9%) | 1317 (71.0%) | 2407 (51.7%) |
| **Sex** |  |  |  |
| Female | 877 (44.7%) | 1247 (67.2%) | 2484 (53.3%) |
| Male | 1087 (55.3%) | 607 (32.7%) | 2169 (46.5%) |
| Missing | 0 (0%) | 1 (0.1%) | 7 (0.2%) |
| **Ethnicity** |  |  |  |
| Asian | 34 (1.7%) | 14 (0.8%) | 106 (2.3%) |
| Black | 1258 (64.1%) | 1593 (85.9%) | 3525 (75.6%) |
| Coloured | 323 (16.4%) | 103 (5.6%) | 317 (6.8%) |
| White | 274 (14.0%) | 74 (4.0%) | 487 (10.5%) |
| Missing | 75 (3.8%) | 71 (3.8%) | 225 (4.8%) |
| **Type of cancer** |  |  |  |
| NADC | 1699 (86.5%) | 697 (37.6%) | 3411 (73.2%) |
| ADC | 206 (10.5%) | 1129 (60.9%) | 1062 (22.8%) |
| Primary Site unknown | 59 (3.0%) | 29 (1.6%) | 187 (4.0%) |
| **ART period** |  |  |  |
| 2004-2007 | 594 (30.2%) | 500 (27.0%) | 2062 (44.2%) |
| 2008-2012 | 822 (41.9%) | 784 (42.3%) | 1647 (35.3%) |
| 2012-2014 | 548 (27.9%) | 571 (30.8%) | 951 (20.4%) |
| **Total** | 1964 (100%) | 1855 (100%) | 4660 (100%) |

NADC=non-AIDS Defining cancer. ADC=AIDS Defining Cancer. ART=Antiretroviral therapy

**Figure 1:**
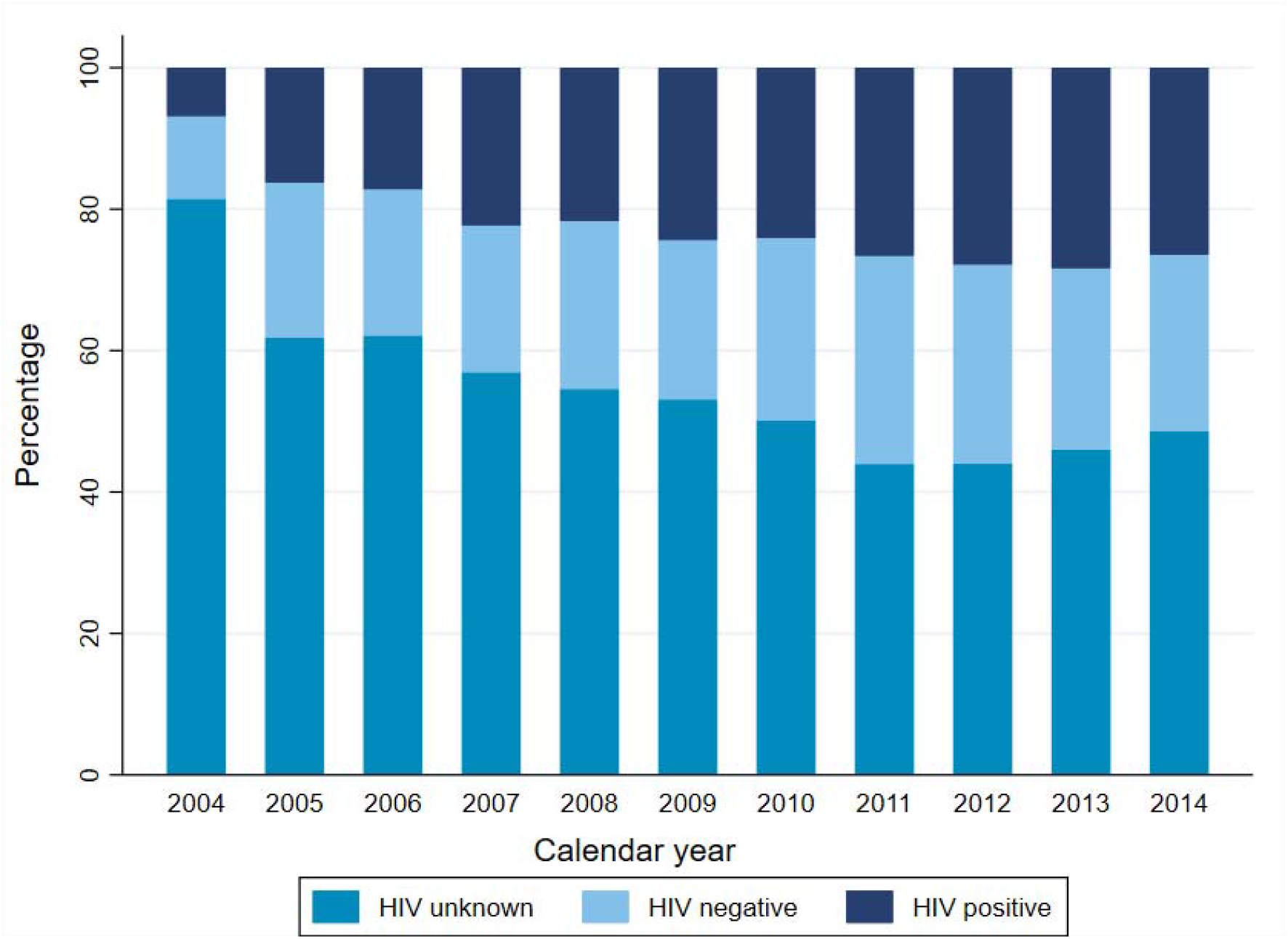
Distribution of HIV unknowns across the study period amongst AYAs with cancer. The trend analysis for proportions was statistically significant across all strata of HIV status (pvalue< 0.001) for all HIV Status relative to the year of cancer diagnosis.

The most frequently diagnosed cancer was Kaposi sarcoma, followed by leukaemia and bone cancer (Figure 2, absolute numbers in Table S2 in supplementary material). Non-AIDS Defining Cancers (NADCs) made up 68% (n=5803) of histologically diagnosed cancers. In AYA with ADCs, 44% (n=1 062) of patients had unknown HIV status vs 59% (n=3 411) of AYA with NADC. The HIV status of 44% (n=617) of AYA diagnosed with Kaposi Sarcoma was unknown, and the HIV status of 43% (n=269) of AYA diagnosed with NHL was unknown (Figure 1). Haematological cancers were the most common cancers in AYAs without HIV: leukaemia was the most frequent diagnosis (n=449), followed by Hodgkin’s lymphoma (n=246), and bone cancers (n=197). In HIV negative AYAs, the top five cancers were similar for male and female participants, but HIV-negative male AYAs had a higher proportion of Hodgkin’s lymphoma and bone cancers.

**Figure 2:**
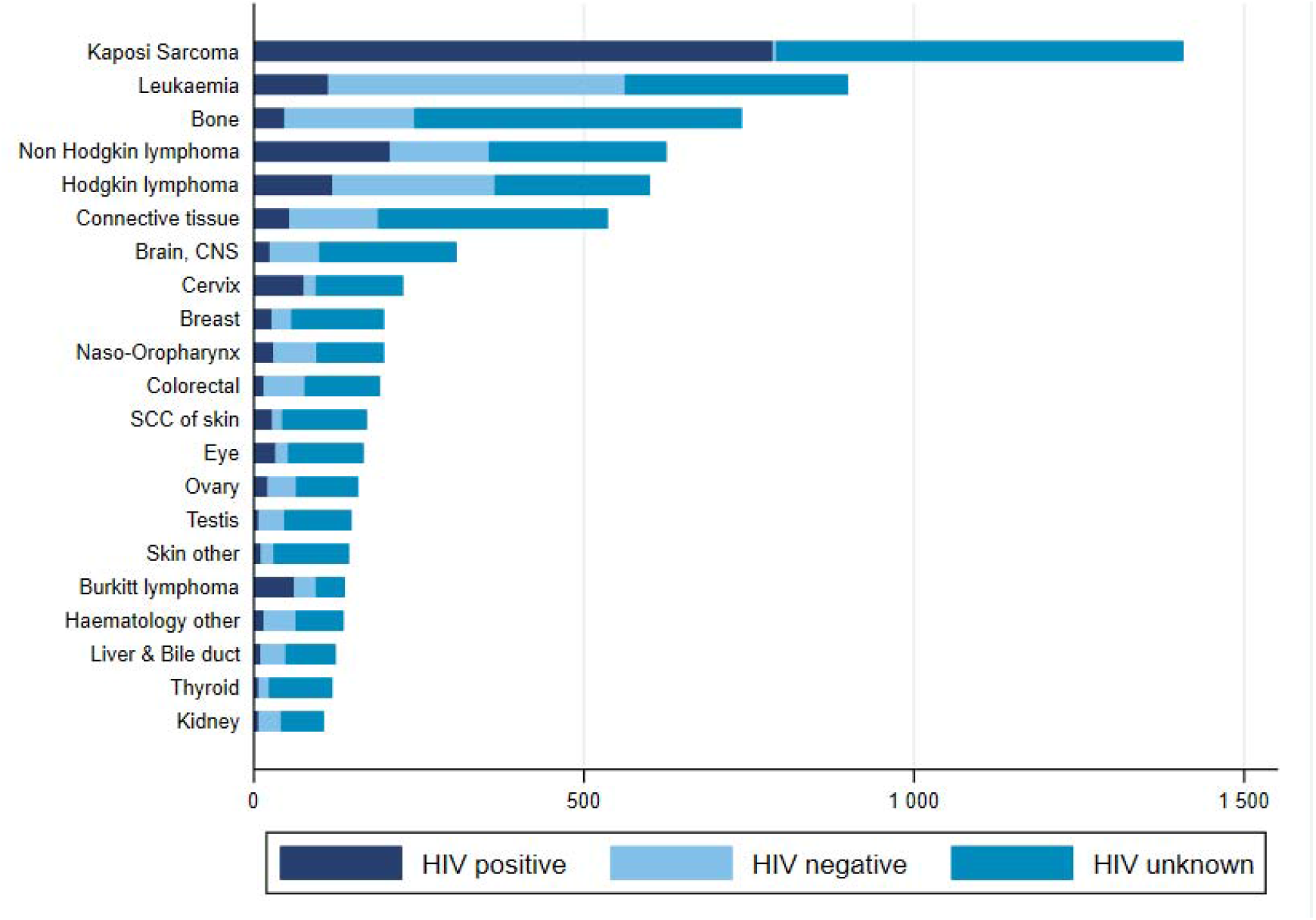
Top 20 cancer in AYAs in the South African public health sector stratified by HIV status. NADC: non-AIDS Defining Cancer. ADC: AIDS Defining Cancer. Brain, CNS: Brain Central Nervous System. SCC of skin: Squamous Cell Carcinoma of skin

Amongst those with recorded HIV status, Kaposi Sarcoma, NHL and Hodgkin’s lymphoma, leukaemia, and cervical cancer were the most frequent cancers in AYAs living with HIV (Figure 1). The top five most frequent cancers amongst female AYAs with HIV were Kaposi sarcoma, NHL, cervical cancer, Hodgkin’s lymphoma, and leukaemia (Figure S1 Supplementary material). For male AYAs with HIV, the most frequently diagnosed cancers were KS, NHL, leukaemia, Hodgkin’s lymphoma, and connective tissue cancers. The proportion of Kaposi sarcoma cases was higher in female AYAs with HIV (71%, n=998) than in male AYAs with HIV (29%, n=409).

The logistic regression analysis revealed higher odds of AIDS Defining Cancers (ADCs) than to NADCs in AYAs with HIV (Table 2). When we compared HIV positive AYAs to HIV negative AYAs, the adjusted odds ratio for AYAs with HIV was 218 (95% CI 89.9–530) for Kaposi sarcoma, 2.18 (95% CI 1.23–3.89) for cervical cancer, and 2.12 (95% CI 1.69–2.66) for NHL. The odds of specific NHL subtypes like Burkitt lymphoma, diffuse Large B-cell lymphoma, and diffuse immunoblastic large B-cell lymphoma were higher in AYAs living with HIV than in AYAs without HIV (Table 2). Anogenital cancers other than cervical cancer were also strongly associated with HIV; adjusted OR was 2.73 (95% CI 1.27–5.86). We did not observe significant odds of leiomyosarcoma in AYAs living with HIV but, of the eight recorded leiomyosarcoma cases with a known HIV result, six were HIV positive and five were female. Odds were not higher for HIV and Hodgkin’s lymphoma or HIV with liver cancer.

**Table 2:** Relationship between HIV and selected cancers amongst AYAs in the South African public health sector.

| Cancer site | Univariable analysis (n=3819) | Multivariable complete case analysis (n=3672) | Multivariable Imputed analysis (n=8103) |
| --- | --- | --- | --- |
|  | Odds ratio (95% CI) | Odds ratio (95% CI) | Odds ratio (95% CI) |
| ADC | 13.3 (11.2-15.8) | 12.0 (9.92-14.5) | 3.15 (2.71-3.65) |
| Kaposi Sarcoma | 288 (119-696) | 218 (89.9-530) | 4.48 (3.64-5.52) |
| NHL | 1.64 (1.34-2.00) | 2.12 (1.69-2.66) | 1.47 (1.17-1.85) |
| Burkitts lymphoma | 1.99 (1.30-3.05) | 2.65 (1.64-4.28) | 2.03 (1.29-3.17) |
| Non-Hodgkin- NOS | 3.31 (1.72-6.37) | 4.28 (2.06-8.92) | 1.75 (1.03-2.99) |
| DLBCL- NOS | 1.52 (1.11-2.09) | 2.03 (1.42-2.90) | 1.49 (1.07-2.10) |
| DILBCL- NOS | 3.61 (1.64-7.97) | 4.75 (2.01-11.3) | 2.25 (1.05-4.82) |
| Mature T-cell- NHL | 0.94 (0.36-2.44) | 1.11 (0.38-3.23) | 0.90 (0.36-2.24) |
| Follicular NHL | 0.79 (0.18-3.55) | 0.93 (0.12-6.89) | 0.84 (0.14-5.08) |
| Cervical cancer | 4.62 (2.75-7.75) | 2.18 (1.23-3.89) | 1.37 (0.92-2.04) |
| NADC | 0.09 (0.08-0.11) | 0.11 (0.09-0.13) | 0.36 (0.32-0.42) |
| Virus-related cancers | 0.56 (0.46-0.68) | 0.64 (0.52-0.80) | 0.80 (0.65-0.98) |
| Anogenital cancers other than cervix | 3.91 (2.00-7.65) | 2.73 (1.27-5.86) | 1.59 (0.77-3.31) |
| Hodgkin's lymphoma | 0.48 (0.38-0.60) | 0.60 (0.47-0.78) | 0.74 (0.58-0.94) |
| Liver & Bile duct | 0.27 (0.14-0.55) | 0.28 (0.13-0.61) | 0.64 (0.33-1.24) |
| Virus-unrelated NADC | 1.88 (1.57-2.25) | 1.69 (1.38-2.07) | 1.30 (1.09-1.56) |
| Bone | 0.23 (0.16-0.32) | 0.29 (0.21-0.42) | 0.70 (0.51-0.93) |
| Brain- CNS | 0.33 (0.21-0.53) | 0.35 (0.20-0.60) | 0.74 (0.53-1.03) |
| Colorectal | 0.25 (0.14-0.44) | 0.15 (0.08-0.28) | 0.51 (0.32-0.79) |
| Connective tissue | 0.41 (0.30-0.57) | 0.44 (0.31-0.64) | 0.72 (0.55-0.96) |
| Eye | 1.85 (1.05-3.27) | 1.11 (0.58-2.13) | 1.15 (0.69-1.91) |
| Haematology | 0.33 (0.18-0.58) | 0.63 (0.34-1.18) | 0.84 (0.52-1.36) |
| Leukaemia | 0.22 (0.18-0.27) | 0.29 (0.23-0.37) | 0.48 (0.39-0.61) |
| Leiomyosarcoma | 3.18 (0.64-15.8) | 2.13 (0.38-11.9) | 1.24 (0.45-3.45) |
| Myeloma | 0.79 (0.18-3.55) | 0.62 (0.09-4.03) | 0.64 (0.13-3.29) |
| Ovary | 0.51 (0.30-0.87) | 0.48 (0.26-0.87) | 0.71 (0.45-1.12) |
| SCC skin | 1.99 (1.06-3.74) | 1.21 (0.60-2.44) | 0.93 (0.56-1.56) |
| Skin | 0.61 (0.29-1.29) | 0.44 (0.19-1.02) | 0.85 (0.50-1.44) |
| Thyroid | 0.46 (0.19-1.12) | 0.29 (0.11-0.77) | 0.85 (0.48-1.49) |
| Testis | 0.19 (0.08-0.42) | 0.28 (0.11-0.68) | 0.71 (0.39-1.31) |

NHL=non-Hodgkin's Lymphoma. DLBCL= Diffuse large B-cell lymphoma. DILBCL= Diffuse immunoblastic large B-cell lymphoma. NOS= Not Otherwise Specified. The multivariable analysis is adjusted for age, sex (were applicable), ethnicity and calendar period. Imputation was done under the missing at random assumption. The variables used to impute for missing HIV status were cancer diagnosis, ethnicity, sex and cancer diagnosis year. The imputed analysis is a multivariable analysis adjusting for age, sex (were applicable), ethnicity and calendar period.

Interaction tests determined that age modified the odds of NHL in AYAs living with HIV; adolescents with HIV had higher odds of NHL (adjusted OR 3.17; 95% CI: 2.35–4.28) than YA with HIV (adjusted OR 1.29; 95% CI 0.93–1.79; p-value for interaction < 0.0001). Ethnicity also modified the odds of Burkitt lymphoma in HIV positive AYAs; Black AYAs with HIV had higher odds of Burkitt lymphoma (adjusted OR 3.84; 95% CI: 2.10–7.04) than non-Black AYAs with HIV (adjusted OR 1.35; 95% CI: 0.43–4.28, p-value for interaction = 0.0199). In the sensitivity analysis that used the imputed dataset multivariable analysis of the imputed dataset, we observed that the, KS (adjusted OR 4.48; 95% CI: 3.64–5.52), cervical cancer (adjusted OR 1.37; 95% CI:0.92–2.04) and anogenital cancers other than cervix (adjusted OR 1.59; 95% CI: 0.77–3.31) had a substantially weaker association or no association. For all other cancers, results were similar to the main analysis of subjects with known HIV status (Table 2).

## DISCUSSION

We observed an association amongst AYAs in the ART era between HIV and ADCs and anogenital cancers other than cervical cancer, including penile, anal, vulvar and vaginal cancers. Among those living with HIV, the proportion of KS was higher in girls and young women than in boys and young men. The combined odds of cancers not associated with HIV were higher in AYAs living with HIV than in those without HIV. We could not ascertain the HIV status of many AYAs diagnosed with HIV related cancers, however, a sensitivity analysis using imputed data yielded qualitatively similar results. We observed higher odds of Burkitt lymphoma black AYAs living with HIV compared to those without HIV and higher odds of NHL in adolescents living with HIV compared to young adults living with HIV.

The record linkage and the additional results determined from the text mining process ensured that we extracted the maximum available HIV results. Our study has several limitations. As in other HIV studies that have used CD4 counts to create HIV cohorts, we assumed that anyone who had a CD4 cell count test was HIV positive. It is possible that CD4 count tests might be performed for other reasons, but we think the risk is low since CD4 tests are usually administered after a positive HIV test. The proportion of patients whose HIV status was unknown might not be representative of the entire HIV population in South Africa because this group includes only those who had laboratory HIV tests. Rapid test results are less likely to appear in the NHLS database (only 10% of cancers had a rapid test result). Our study shares the same limitations as the proportionate incidence analysis. Since out study population included only AYAs with cancer just like in proportionate incidence analysis, the odds ratios may have been overestimated. For the most common cancers, the odds ratio might reflect how frequently a cancer is observed and not the actual strength of association between HIV and the cancer. Using all other cancers as a comparison group may have also led to underestimating the strength of the association, especially for cancers with overlapping risk factors. However, this does not necessarily mean that the effects of the last two limitations cancel out.

It is known that the risk of ADCs is higher in AYALHIV.[10–12,14,24,25] In our study, KS was the cancer most strongly associated with HIV. HIV cohort studies have reported increased KS incidence among children and adolescents under 16.[10,12] A multicohort study found KS risk was higher in HIV positive adolescents and children from Southern Africa than in the same age group in other regions of the world.[26] In South Africa, where treatment and retention in care rates for AYAs with HIV are low[3], poorly controlled HIV infection amongst AYAs may increase the odds of KS. The South African National HIV Prevalence Survey of 2017 revealed that about 60% of young adults (ages 15–24) living with HIV were not on ART.[27] Untreated AYALHIV are likely to develop immunodeficiency which increases their risk of developing KS.[26]

The risk of cervical cancer in this young adult population may be increased for several reasons. In South Africa in 2017, girls and young women were much more likely to be HIV positive (10.9% prevalence) than boys and young men (4.8%).[27] Biological factors may account for higher HIV prevalence in girls and young women, along with socio-economic factors that encourage risky sexual behaviour including transactional and intergenerational sexual relationships.[27] High prevalence and poorly controlled HIV can increase the risk of HPV co-infection in an age group less likely to be screened for cervical cancer, which in turn increases cervical cancer risk and risk of other anogenital cancers amongst AYAs. A study in the Western Cape province of South Africa found AYALHIV had higher HPV prevalence and more HPV subtypes than AYAs without HIV.[8] In contrast to other studies on cancer in AYALHIV, we observed three cervical cancer cases in AYAs between 14 and 16; two of these young women were HIV positive. HIV cohorts in South Africa and the USA have not identified cervical cancer in children and adolescents under 16,[10,14] but cervical cancer risk and incidence has been on the increase in the ART era for those between 18–24.[28] Early sexual debut, and subsequent early exposure to causative agents like HPV may explain this early presentation with cervical cancer in South Africa[8,29,30], but more studies are needed to explore this phenomenon.

Lymphomas are often misdiagnosed as tuberculosis in people living with HIV in our setting, slowing diagnosis and worsening the prognosis.[31] This might explain the significantly lower odds of Hodgkin lymphoma, a cancer associated with HIV in our study population. Like other studies, we found non-Hodgkin’s lymphoma (NHL) was associated with HIV.[10,14] NHL is associated with poor adherence to ART and low rates of viral suppression, and NHL risk is high in HIV positive individuals on ART even when their disease is controlled.[14,32,33] This may be because HIV activates the CD40 receptors on B-cells like Epstein Barr virus (EBV) would in EBV related cancers such as Burkitt lymphoma.[32] We expect poor ART coverage and retention in care among AYAs with HIV increases this risk, but researchers still need to determine NHL risk in virally suppressed and non-suppressed patients in our setting. From, the interaction analysis, the odds of NHL were higher in adolescents with HIV compared to young adults with HIV. This observation could be as a result of the predominance of lymphoblastic and Burkitt lymphoma, which are more common in younger ages.[34] We also found that the odds of Burkitt lymphoma were higher in HIV positive black AYAs compared to the Other ethnicities. Burkitt lymphoma in South Africa is more likely to be found in white children aged 0–14 years than in Black children[22], but this is a different age group to that in our study. We recommend physicians maintain a high suspicion index for lymphomas in AYALHIV and take care not to misdiagnose them as tuberculosis, thereby delaying care.

Other studies identified an association between leiomyosarcoma and HIVs.[13] Although not statistically significant, the odds of leiomysorcoma were higher in AYALHIV than in AYAs without HIV. Since leiomyosarcoma is rare, the association between leiomyosarcoma and HIV needs further study. Likewise, after we adjusted for the interaction of HIV with age and calendar period, AYALHIV had an increased risk of connective tissue cancer, but this finding did not reach statistical significance.

Although the proportion of subjects with unknown HIV status decreased with time, HIV testing for HIV related cancers remained low. The HIV status of many AYAs with KS, cervical cancer and NHL was unknown. In South Africa, HIV testing uptake is lower in AYAs than in adults[27] and is mostly opportunistic.[35] An AYA is most likely to be tested if they present to a health care facility with symptoms linked to a sexually transmitted infection or if a female AYA visits a reproductive health clinic.[36] Known HIV results were dependent on cancer type. The results from the complete case analysis were mostly overestimated when compared to the imputed analysis thus pointing towards differential misclassification of HIV results.

Because AYALHIV are at higher risk of ADCs and anogenital cancers and many AYAs with HIV-related cancers are not tested for HIV, HIV programmes for AYAs should extend testing coverage, link AYAs to care, and make sure to retain them. AYALHIV have a higher risk of cervical and other anogenital cancers because of the high frequency of HPV co-infection, exacerbated by sexual debut and young age. We recommend sexually active AYALHIV be screened for cervical cancer at HIV diagnosis and followed up frequently, as per the new cervical cancer guidelines so that potential CC can be identified early and treated. South Africa has already introduced an HPV vaccination programme for nine-year-old girls, and this program should be extended to AYALHIV. Those not yet infected with HPV sub-types covered by the vaccines should be vaccinated, regardless of age.

## Conclusion

This is the first nationwide study in South Africa to compare the distribution of cancers in adolescents and young adults (AYAs) by HIV status. AIDS defining cancers (ADCs) and anogenital cancers other than cervix cancer were more common in HIV-positive than in HIV-negative AYAs. AYAs with cancer are a key population for HIV testing, however this study showed that many AYAs with ADCs are not tested for HIV. Targeted HIV testing for AYAs should be followed by the immediate start of ART after a positive diagnosis, accompanied by cervical cancer screening and vaccination against HPV to decrease cancer burden in adolescents and young adults living with HIV in South Africa.

## Data Availability

The datasets used and/or analysed during the current study are available from the corresponding author upon reasonable request.

## Authors’ contributions

ME, ES, MS and JB contributed towards the study design. TD contributed towards literature search, data analysis and drafting of first version of manuscript. ES and MS contributed towards data acquisition. AS contributed towards data linkage. VO contributed towards text mining of cancer pathology reports to assign HIV status. All authors contributed towards data interpretation and critical comments on the first and subsequent drafts of the manuscript. All authors read and approved the final manuscript.

## Ethics and dissemination

Permission to use the routinely collected NHLS and NCR data was sought from the relevant authorities. Ethical approval to conduct the study was granted by the University of the Witwatersrand Human Research Ethics Committee [Ethics certificate numbers (SAM: M160944) and (BCAH: M171083)].

## Patient and public involvement

Individual consent was not required but permission to use the routinely collected National Health Laboratory Service and National Cancer Registry data was sought and approved by the relevant authorities.

## Patient consent for publication

Not required.

## Acknowledgements

The authors would like to thank all funders, the University of the Witwatersrand, the National Health Laboratory Service (NHLS), the NHLS’s Corporate Data Warehouse (special thanks to Sue Candy) and the National Cancer Registry. We would also like to thank Kali Tali for her editorial input.

---

**Online Supplement Table S1:** Distribution of AYAs with known and unknown HIV status by characteristics used in the imputation model.

| **Characteristics** | **Unknown HIV status n (%)** | **Known HIV status n (%)** |
| --- | --- | --- |
| **Age [years]** |  |  |
| 10 | 162 (47.1%) | 182 (52.9%) |
| 11 | 152 (46.8%) | 173 (53.2%) |
| 12 | 160 (48.8%) | 168 (51.2%) |
| 13 | 190 (52.9%) | 169 (47.1%) |
| 14 | 210 (65%) | 113 (35%) |
| 15 | 210 (56.3%) | 163 (43.7%) |
| 16 | 233 (61.5%) | 146 (38.5%) |
| 17 | 259 (61.4%) | 163 (38.6%) |
| 18 | 276 (57.4%) | 205 (42.6%) |
| 19 | 284 (57.3%) | 212 (42.7%) |
| 20 | 337 (56.5%) | 259 (43.5%) |
| 21 | 401 (57.9%) | 291 (42.1%) |
| 22 | 451 (53.6%) | 391 (46.4%) |
| 23 | 530 (53.2%) | 467 (46.8%) |
| 24 | 576 (50.3%) | 570 (49.7%) |
| **Sex** |  |  |
| Female | 2374 (53.6%) | 2051 (46.4%) |
| Male | 2057 (55.9%) | 1621 (44.1%) |
| **Ethnicity** |  |  |
| Asian | 106 (68.8%) | 48 (31.2%) |
| Black | 3521 (55.3%) | 2850 (44.7%) |
| Coloured | 317 (42.7%) | 426 (57.3%) |
| White | 487 (58.3%) | 348 (41.7%) |
| **Cancer diagnosis date** |  |  |
| 2004 | 575 (81%) | 135 (19%) |
| 2005 | 415 (61.8%) | 257 (38.2%) |
| 2006 | 492 (62.8%) | 292 (37.2%) |
| 2007 | 458 (57.1%) | 344 (42.9%) |
| 2008 | 443 (54.2%) | 375 (45.8%) |
| 2009 | 438 (53.3%) | 383 (46.7%) |
| 2010 | 369 (50%) | 369 (50%) |
| 2011 | 327 (43.6%) | 423 (56.4%) |
| 2012 | 348 (42.9%) | 463 (57.1%) |
| 2013 | 283 (46.2%) | 329 (53.8%) |
| 2014 | 283 (48.4%) | 302 (51.6%) |
| **Cancer site** |  |  |
| Anus | 13 (56.5%) | 10 (43.5%) |
| BCC | 60 (85.7%) | 10 (14.3%) |
| Bladder | 18 (85.7%) | 3 (14.3%) |
| Bone | 467 (66.3%) | 237 (33.7%) |
| Brain, CNS | 192 (67.8%) | 91 (32.2%) |
| Breast | 137 (71%) | 56 (29%) |
| Burkitt lymphoma | 40 (30.5%) | 91 (69.5%) |
| Cervix | 128 (57.9%) | 93 (42.1%) |
| Colorectal | 108 (59.7%) | 73 (40.3%) |
| Connective tissue | 334 (65.1%) | 179 (34.9%) |
| Endocrine | 22 (42.3%) | 30 (57.7%) |
| Eye | 109 (69.4%) | 48 (30.6%) |
| Gum | 1 (100%) | 0 (0%) |
| Haematology other | 68 (52.7%) | 61 (47.3%) |
| Hodgkin lymphoma | 228 (39.8%) | 345 (60.2%) |
| Ill defined | 7 (100%) | 0 (0%) |
| Kaposi Sarcoma | 597 (43.9%) | 764 (56.1%) |
| Kidney | 63 (61.2%) | 40 (38.8%) |
| Larynx | 1 (25%) | 3 (75%) |
| Leukaemia | 321 (37.3%) | 539 (62.7%) |
| Lip | 8 (61.5%) | 5 (38.5%) |
| Liver & Bile duct | 70 (61.4%) | 44 (38.6%) |
| Lung | 23 (76.7%) | 7 (23.3%) |
| Melanoma | 76 (81.7%) | 17 (18.3%) |
| Mesothelioma | 3 (100%) | 0 (0%) |
| Mouth | 32 (60.4%) | 21 (39.6%) |
| Myeloma | 5 (45.5%) | 6 (54.5%) |
| Naso-Oropharynx | 95 (51.1%) | 91 (48.9%) |
| Non Hodgkin lymphoma | 253 (42.2%) | 347 (57.8%) |
| Oesophagus | 14 (66.7%) | 7 (33.3%) |
| Other specified | 42 (57.5%) | 31 (42.5%) |
| Ovary | 90 (59.6%) | 61 (40.4%) |
| Pancreas | 4 (57.1%) | 3 (42.9%) |
| Penis | 3 (50%) | 3 (50%) |
| Placenta | 53 (65.4%) | 28 (34.6%) |
| Primary site unknown | 172 (66.9%) | 85 (33.1%) |
| Prostate | 15 (83.3%) | 3 (16.7%) |
| SCC of skin | 120 (74.1%) | 42 (25.9%) |
| Salivary gland | 51 (68%) | 24 (32%) |
| Skin other | 108 (78.8%) | 29 (21.2%) |
| Small intestine | 7 (77.8%) | 2 (22.2%) |
| Stomach | 24 (58.5%) | 17 (41.5%) |
| Testis | 98 (69%) | 44 (31%) |
| Thyroid | 89 (80.2%) | 22 (19.8%) |
| Tongue | 7 (77.8%) | 2 (22.2%) |
| Uterus | 28 (57.1%) | 21 (42.9%) |
| Vagina | 10 (50%) | 10 (50%) |
| Vulva | 17 (38.6%) | 27 (61.4%) |

**Online Supplement Table S2:** Top 20 cancer in AYAs in the South African public health sector stratified by HIV status.

| **Cancer Site** | **HIV positive** | **HIV negative** | **HIV unknown** |
| --- | --- | --- | --- |
| NADC | 697 | 1699 | 3411 |
| ADC | 1129 | 206 | 1062 |
| Kaposi Sarcoma | 786 | 5 | 617 |
| Leukaemia | 113 | 449 | 338 |
| Bone | 46 | 197 | 496 |
| NHL | 206 | 150 | 269 |
| Hodgkin lymphoma | 119 | 246 | 235 |
| Connective tissue | 54 | 134 | 348 |
| Brain, CNS | 24 | 75 | 208 |
| Cervix | 76 | 18 | 132 |
| Breast | 27 | 30 | 140 |
| Naso-oropharynx | 30 | 65 | 102 |
| Colorectal | 15 | 62 | 114 |
| SCC of Skin | 28 | 15 | 128 |
| Eye | 33 | 19 | 114 |
| Ovary | 21 | 43 | 94 |
| Testis | 7 | 39 | 102 |
| Skin Other | 11 | 19 | 114 |
| Haematology other | 15 | 48 | 73 |
| Liver and Bile duct | 10 | 38 | 76 |
| Thyroid | 7 | 16 | 96 |
| Kidney | 7 | 34 | 65 |

**Figure S3:**
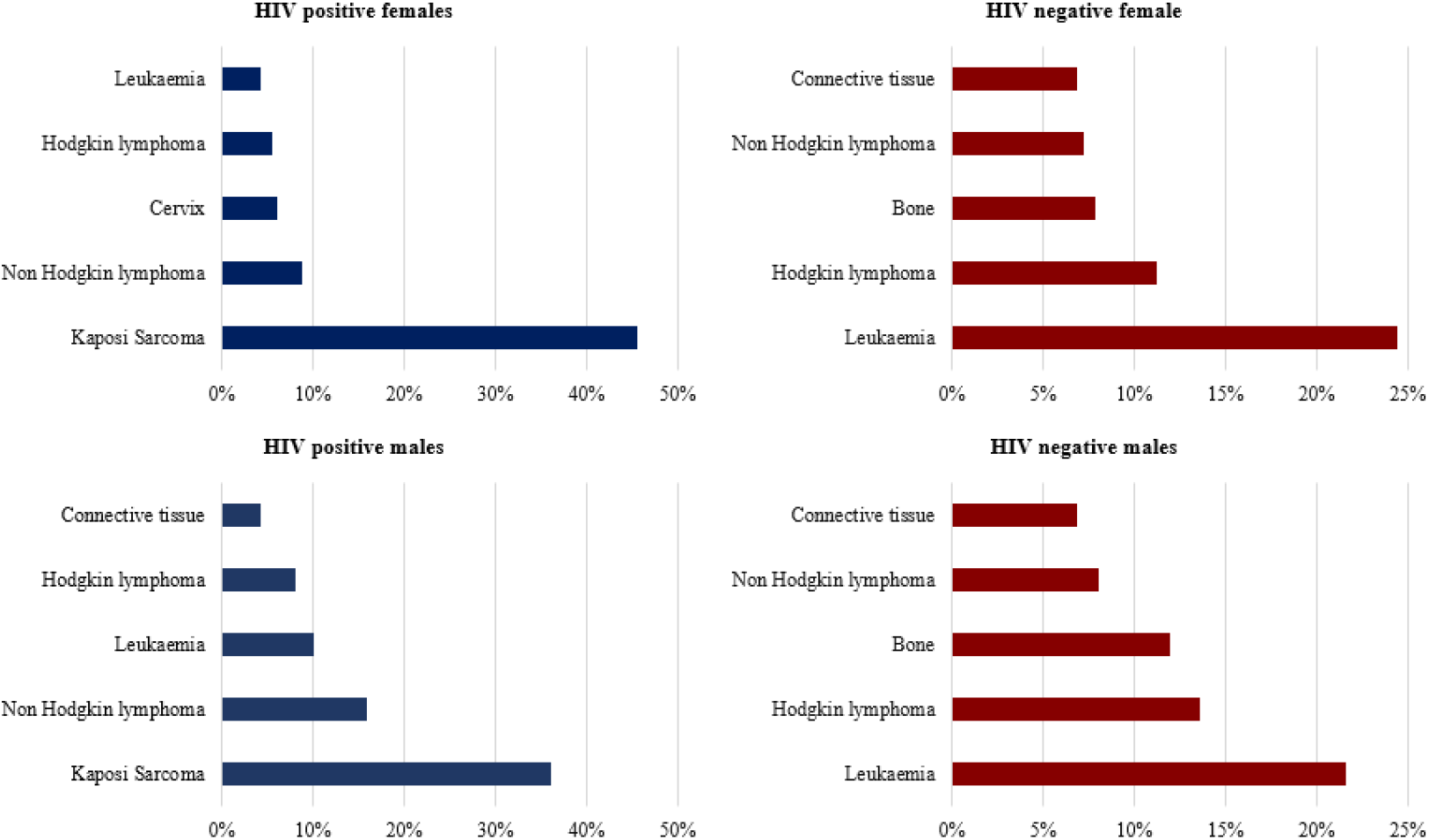
Top five cancers in AYAs stratified by sex and HIV status

